# Blood Pressure and Cerebral Oxygenation with Physiologically-based Cord Clamping: A Sub-Study of the BabyDUCC Trial

**DOI:** 10.1101/2023.09.19.23295703

**Authors:** Shiraz Badurdeen, Douglas A Blank, Monsurul Hoq, Flora Y Wong, Calum T Roberts, Stuart B Hooper, Graeme R Polglase, Peter G Davis

**Affiliations:** Newborn Research Centre, The Royal Women’s Hospital, Melbourne VIC 3052, Australia; The Ritchie Centre, Hudson Institute of Medical Research, Clayton VIC 3168, Australia; Department of Paediatrics, Mercy Hospital for Women, Heidelberg VIC 3084, Australia; Monash Newborn, Monash Children’s Hospital, Clayton VIC 3168, Australia; Department of Paediatrics, Monash University, Clayton VIC 3168, Australia; Clinical Epidemiology and Biostatistics Unit and Clinical Sciences Research, Murdoch Children’s Research Institute, Melbourne VIC 3052, Australia; Department of Obstetrics and Gynaecology, Monash University, Wellington Rd, Clayton VIC 3800, Australia; Department of Obstetrics and Gynaecology, The University of Melbourne, Parkville VIC 3052, Australia

**Author notes:** Corresponding Author: Shiraz Badurdeen, Address: Newborn Research Centre, The Royal Women’s Hospital, Melbourne, VIC 3052, Australia. Joint senior authors (Graeme R Polglase, Peter G Davis).

**Keywords:** Neonate, resuscitation, cord clamping, blood pressure, cerebral oxygenation

## Abstract

**Background:** Cord-clamping strategies may modify blood pressure (BP) and cerebral tissue oxygen saturation (rStO_2_) immediately after birth.

**Methods:** We conducted a sub-study nested within the Baby-Directed Umbilical Cord-Clamping trial. Infants ≥32^+0^ weeks’ gestation assessed as requiring resuscitation were randomly allocated to either physiologically-based cord clamping (PBCC), where resuscitation commenced prior to umbilical cord clamping, or standard care where cord clamping occurred early (ECC). In this single-site sub-study, we obtained additional measurements of pre-ductal BP and rStO_2_. In a separate observational arm, non-randomised vigorous infants received 2 minutes of deferred cord clamping (DCC) and contributed data for reference percentiles.

**Results:** Among 161 included infants, n=55 were randomly allocated to PBCC (n= 30) or ECC (n= 25). The mean (SD) BP at 3-4 minutes after birth (primary outcome) in the PBCC group was 64 (10) mmHg compared to 62 (10) mmHg in the ECC group, mean difference 2 mmHg (95% confidence interval −3 – 8 mmHg, p= 0.42). BP and rStO_2_ were similar across both randomised arms and the observational arm (n=106).

**Conclusion:** We found no difference in BP or rStO_2_ with the different cord clamping strategies. We report reference ranges for BP and rStO_2_ for late-preterm and full-term infants receiving DCC.

**Impact:**

- Among late-preterm and full-term infants receiving resuscitation at birth, blood pressure (BP, at 3-4 minutes and 6-7 minutes) and cerebral tissue oxygen saturation (rStO_2_) are not influenced by timing of cord clamping in relation to establishment of ventilation.
- Infants in this study did not require advanced resuscitation, where different cord clamping strategies may yet influence BP and rStO_2_.
- The reference ranges for BP and rStO_2_ represent the first, to our knowledge, for vigorous late-preterm and full-term infants receiving deferred cord clamping.
- rStO_2_ >90% (∼90^th^ percentile) may be used to define cerebral hyperoxia, for instance when studying oxygen supplementation after birth.

## Introduction

Fetal compromise leads to haemodynamic instability during the transitional period after birth.^1^ The establishment of ventilation prior to umbilical cord clamping (UCC) provides greater haemodynamic stability in sheep models of perinatal asphyxia and preterm birth.^2–5^ However, the impact of the relationship between onset of ventilation and timing of UCC on haemodynamic stability in late-preterm and term infants receiving resuscitation is not known.

In asphyxiated lambs, UCC prior to ventilation resulted in rebound hypertension within 5 minutes of resuscitation and was associated with markers of cerebral injury, including protein extravasation in the subcortical and periventricular regions.^2^ Resuscitating lambs with the umbilical cord intact greatly mitigated the rebound hypertension and brain injury.^2^ This was likely due to the low-resistance placental vascular bed within the systemic circulation during the recovery in cardiac function.

A further mechanism of secondary brain injury may be the prolongation of cerebral hypoxia. Studies in preterm lambs found that UCC prior to ventilation results in a decrease in arterial oxygen saturation and cerebral oxygenation.^4^ By providing continued return of oxygenated blood to the left side of the heart, delaying cord clamping until lung aeration improved both systemic and cerebral oxygenation.^5^ Conversely, in asphyxiated animals, cerebral hyperoxia occurs following the rebound surge in cerebral blood flow and use of supplemental oxygen during resuscitation.^6^ Cerebral hyperoxia leads to oxidative stress and mitochondrial dysfunction, potentially exacerbating cellular injury from the primary asphyxial insult.^7–9^

Pulse-oximetry to monitor arterial oxygen saturation (SpO_2_) provides little information about the adequacy of cerebral oxygen delivery as it does not account for cerebral blood flow, haemoglobin concentration or metabolic demand.^6,10^ Continuous monitoring of regional cerebral tissue oxygen saturation (rStO_2_) and fractional tissue oxygen extraction (cFTOE) using near-infrared spectroscopy (NIRS) has been performed during fetal-to-neonatal transition.^11,12^ By measuring predominantly venous oxygen saturation, this technique enables an estimation of the balance between cerebral oxygen delivery and consumption.^13^ Previous studies describing normal ranges of rStO_2_ and cFTOE during full-term neonatal transition have focused on infants with early cord clamping (ECC).^14,15^ However, the reported values may not apply to infants who have deferred cord clamping (DCC), which is now the recommended standard care for vigorous term infants.^16^

We aimed to evaluate whether establishment of ventilation before UCC modifies systemic BP and cerebral oxygen saturation during the transitional period in late preterm and term infants who require resuscitation at birth. We also aimed to establish normal reference ranges from infants who were at-risk of requiring resuscitation but transitioned normally and received DCC. We hypothesised that UCC prior to ventilation results in increased BP and decreased cerebral oxygenation when compared to infants who establish ventilation prior to UCC.

## Methods

This was a sub-study nested in the Baby-DUCC randomised controlled trial (RCT) and cohort study at the Royal Women’s Hospital (RWH) site.^17^ Approval was obtained from the RWH Human Research Ethics Committee (RWH ethics 17/19). Antenatal consent was obtained from parents in addition to consent for the Baby-DUCC trial. Recruitment was between March 2019 and April 2021. This study was prospectively registered: ACTRN12619000277145.

### Participants

Infants were eligible for participation in the Baby-DUCC RCT and cohort study if they fulfilled the following inclusion criteria: ≥32^+0^ weeks’ gestation at birth, paediatric doctor requested to attend an at-risk birth, researcher present. Infants with any of the following criteria were excluded: known congenital anomalies compromising cardiorespiratory transition, high risk of obstetric complications requiring early cord clamping, monochorionic twins and multiples >2.

For this study, additional inclusion criteria applied: availability of additional research resources to measure blood pressure (BP) and cerebral oxygenation, antenatal consent for the additional study measurements.

### Procedures

In the BabyDUCC RCT, infants assessed as requiring resuscitation within 1 minute of birth were randomly allocated 1:1 to either standard care, where cord clamping occurred early (ECC) prior to resuscitation, or physiologically based cord clamping (PBCC).^18^ With PBCC, resuscitation commenced and respiratory support, if needed, was provided prior to UCC. Infants in the PBCC group receiving positive pressure ventilation had UCC deferred until ≥2 minutes after birth and until ≥60 seconds after exhaled carbon dioxide was detected on a disposable colorimetric carbon dioxide detector (Pedicap, Medtronic, USA) placed between the facemask and T-Piece. Infants in the PBCC group who breathed without positive pressure ventilation had UCC at ≥2 minutes after birth. This was to ensure that the pulmonary circulation was established prior to cord clamping. Infants in the ECC group had UCC immediately after randomisation and were transferred to a resuscitation trolley prior to commencing respiratory support.

Infants who were vigorous immediately after birth were not randomised and received 2 minutes of deferred cord clamping (DCC). These infants were eligible for inclusion in the observational study arm. Non-randomised infants who went on to receive respiratory support in the delivery room were excluded.^19^

The decision to provide resuscitation and the type of support provided were at the discretion of the attending first-line doctor trained in the Australian and New Zealand Committee on Resuscitation Neonatal Resuscitation Guidelines.^20,21^ Respiratory support was commenced with a Giraffe stand-alone resuscitation system (GE Healthcare, United States of America) set to pressures of 30/5 cmH_2_O in 21% FiO_2_.

### Measured Outcomes

The pre-specified primary outcome for this sub-study was mean BP at 3-4 minutes after birth measured on the right upper arm (Neonatal Single-Patient Non-Invasive Blood Pressure Cuff, sizes 3 and 4, Philips USA). Secondary outcomes were mean BP at 6-7 minutes after birth, systolic and diastolic BP at both timepoints, change in cerebral oxygenation (rStO_2_ and cFTOE) over the 10 minutes after birth, and rStO_2_ at 1 hour after birth.

### Data acquisition

Immediately after birth, a researcher dried the infant and placed three ECG chest leads and a preductal pulse oximeter to monitor the infant’s heart rate (HR) and SpO_2_. At 3-4 min and 6-7 min after birth, pre-ductal BP was measured using a Non-Invasive BP cuff (size 4 for term infants, size 3 for preterm infants, Philips Healthcare, USA) at the right upper arm. A neonatal NIRS sensor (8004CB-NA, SenSmart X-100, Nonin, USA) was placed on the right forehead and secured/protected from ambient light with a hat. HR, SpO_2_ and BP were displayed on a portable Intellivue X2 (Philips Healthcare, USA) and cerebral oxygen saturation displayed on a portable SenSmart X-100 monitor, visible to the clinician. A GoPro Hero Session (GoPro, USA) camera captured the monitor screens, T-piece manometer, oxygen blender dial and audio of the events after birth. The videos were downloaded for offline manual data extracted to ensure high fidelity.

Blinded data extraction was performed for randomised infants. The video recording was cropped to include only the monitor screens and muted to sound before being shared with an off-site researcher. For all infants, HR and SpO_2_ data were extracted every 10 s and rStO_2_ from NIRS every 20 s until 10 min after birth. SpO_2_ readings were only accepted if plethysmograph waveforms showed adequate signal quality. rStO_2_ readings were only accepted if there was no signal interference error shown and no rapid fluctuation suggesting loss of contact. A spot rStO_2_ reading was taken at 1h after birth if it did not interfere with skin-to-skin contact and breastfeeding. This reading was unblinded to group allocation.

### Statistical Analysis

Based on previous studies,^22,23^ we estimated that infants in the ECC group would have a mean (SD) BP of 55(10) mmHg. To detect a mean difference of 10 mmHg between study groups, accepting a 2-sided alpha of 0.05 and 90% power, we calculated a sample size of 24 infants per group. To accommodate 10% attrition rate for detecting the primary outcome due to monitoring failure, we increased our total sample size to 26 infants in each group (n=52).

All analyses were specified *a priori* based on intention-to-treat. For the primary outcome, we calculated the difference in group means, 95% confidence interval (CI) and p-value from an independent samples t-test. Subgroup analyses were planned for the randomisation strata of preterm (32^+0^-35^+6^ weeks’ gestation), non-emergency birth ≥36^+0^ weeks’ gestation, and emergency birth ≥36^+0^ weeks’ gestation. Emergency births were defined as instrumental and unplanned caesarean births.

Data from non-randomised infants who received ≥2 min DCC per protocol and remained vigorous after cord clamping (no respiratory support) until ≥10 min after birth were used to determine reference ranges of pre-ductal BP at 3-4 min and 6-7 min, as well as reference percentiles of rStO_2_ and cFTOE. For rStO_2_ and cFTOE, we used methodology originally proposed by Royston and recommended by Cole for longitudinal data.^17,24,25^ This involved fitting nonlinear regression models to the mean using fractional polynomials in minutes after birth. Mean values were estimated using mixed-effect regression using the same power variables of minutes after birth as fixed effects, as well as infant and time as random effects. Type of birth and interaction between time after birth and type of birth as covariates were included in the model based on the data and previous literature.^17,26^ The percentiles were then calculated by adding or subtracting standard deviation of values multiplied by z-scores based on standard normal distribution to the mean values.

## Results

One-hundred and ninety-two infants were enrolled. After birth, 55 infants were randomly allocated to the PBCC (n= 30) or ECC (n= 25) arms. Among 137 non-randomised infants, 106 were included in the observational study arm (Figure 1); infants born between 32^+0^ – 34^+6^ we excluded due to small numbers (n=2), as were infants who received respiratory support after DCC (n=23), infants who did not complete 2 min of DCC (n=5) and one infant where we were unable to obtain data.

**Fig 1.**
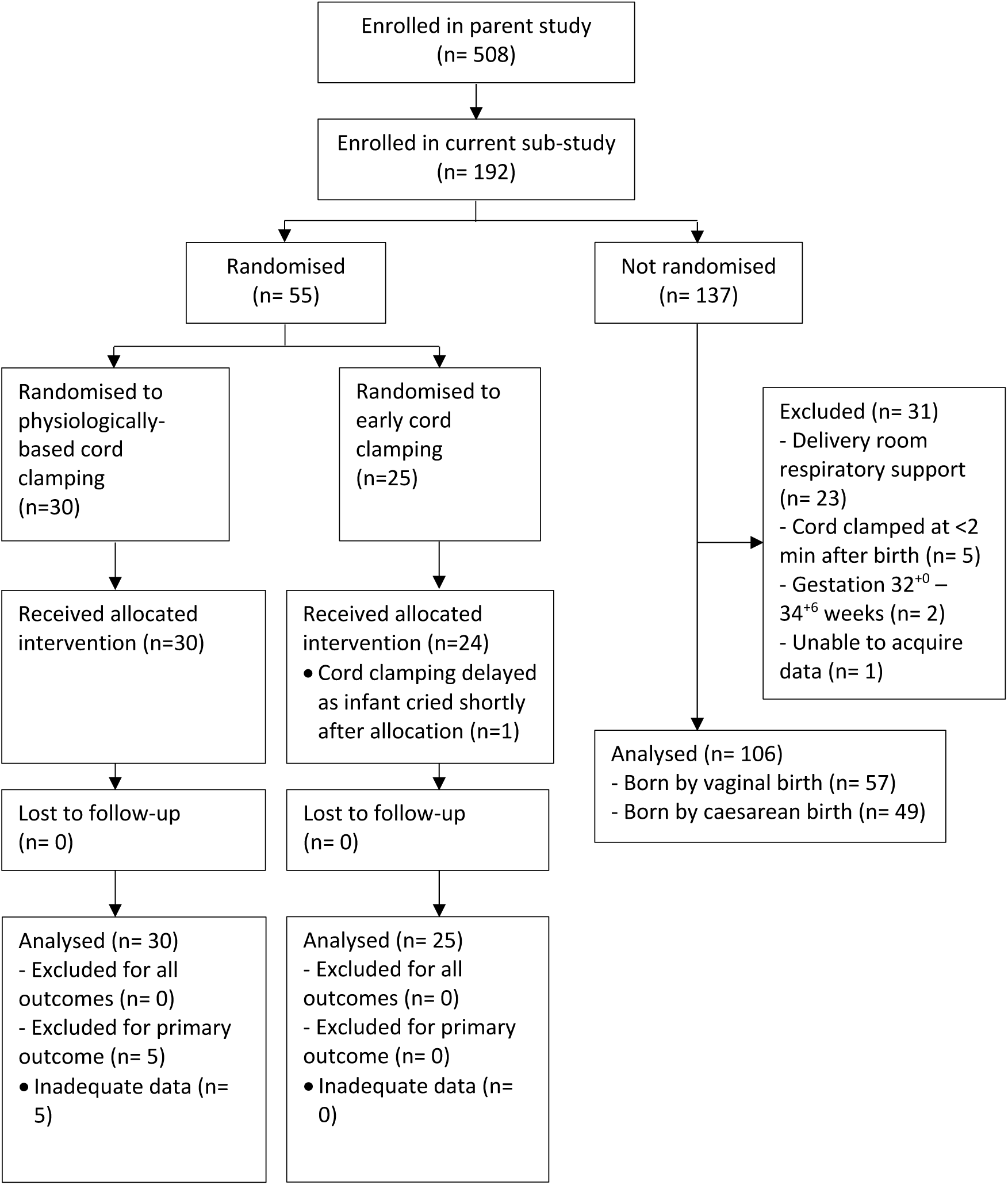
Flow diagram of participants in the study.

The baseline characteristics reflect the high-risk population enrolled (Table 1). Aside from a larger proportion of mothers in the PBCC arm having a medical complication of pregnancy compared to the ECC arm (40% versus 16%), the groups were similar. The median gestation at birth was 39+2 weeks in both randomised arms.

**Table 1.** Baseline characteristics of included participants.

|  | **PBCC**<br>**arm**<br>**(N=30)** | **ECC**<br>**arm**<br>**(N=25)** | **Observational**<br>**arm**<br>**(N=106)** |
| --- | --- | --- | --- |
| **Maternal** |  |  |  |
| Age (years), median [IQR] | 33 [30-35] | 31 [29-33] | 33 [29-35] |
| Primiparity | 27 (90%) | 22 (88%) | 88 (83%) |
| Any medical complication of pregnancy<sup>a</sup> | 12 (40%) | 4 (16%) | 24 (23%) |
| Spontaneous onset of labour | 8 (27%) | 6 (24%) | 34 (32%) |
| Antenatal oxytocin infusion | 18 (60%) | 16 (64%) | 35 (33%) |
| Analgesia/anaesthesia |  |  |  |
| - Spinal or epidural | 30 (100%) | 25 (100%) | 100 (94%) |
| - Other | 0 (0%) | 0 (0%) | 6 (6%) |
| **Birth** |  |  |  |
| Reason for paediatric attendance |  |  |  |
| - Preterm < 37<sup>+0</sup> weeks | 5 (17%) | 3 (12%) | 8 (8%) |
| - Fetal growth restriction | 1 (3%) | 0 (0%) | 4 (4%) |
| - Meconium-stained liquor | 5 (17%) | 6 (24%) | 34 (32%) |
| - Abnormal CTG | 11 (37%) | 10 (40%) | 62 (58%) |
| - Breech/transverse lie | 13 (43%) | 12 (48%) | 18 (17%) |
| - Instrumental birth | 11 (37%) | 10 (40%) | 47 (44%) |
| - Unplanned caesarean section | 7 (23%) | 5 (20%) | 33 (31%) |
| Labour complications |  |  |  |
| - Failure to progress | 1 (3%) | 0 (0%) | 14 (13%) |
| - Prolonged 2<sup>nd</sup> stage<sup>b</sup> | 4 (13%) | 6 (24%) | 30 (28%) |
| - Difficult extraction | 19 (63%) | 15 (60%) | 23 (22%) |
| - None | 9 (30%) | 6 (24%) | 44 (42%) |
| Gestational age (weeks), median [IQR] | 39.3 [38.4-40.7] | 39.3 [38.3-40.0] | 39.6 [38.9-40.7] |
| Birthweight (kg), median [IQR] | 3.42 [3.17-3.81] | 3.39 [2.89-3.64] | 3.43 [3.14-3.75] |
| Female sex | 18 (60%) | 14 (56%) | 54 (55%) |
| Time from birth (s) to |  |  |  |
| - Randomisation, median [IQR] | 23.5 [20-34] | 21.0 [16-29] | N/A |
| - Cord clamping, median [IQR] | 136 [125-150] | 36 [26-44] | 129 [123-141] |
| - Maternal oxytocin administration, median [IQR] | 138 [130-147] | 51 [42-61] | 130 [125-144] |
| **Resuscitation** |  |  |  |
| Any resuscitation | 30 (100%) | 19 (76%) | 0 (0%) |
| - Stimulation alone | 10 (33%) | 5 (26%) |  |
| - CPAP (+/- oxygen) | 4 (13%) | 4 (21%) |  |
| - PPV (mask) | 16 (53%) | 10 (53%) |  |
| Oxygen supplementation |  |  |  |
| - Any oxygen given | 12 (40%) | 11 (44%) | 0 (0%) |
| - Initial FiO<sub>2</sub>, median [IQR] | 0.40 [0.30-0.50] | 0.30 [0.30-0.45] |  |
| - Maximum FiO<sub>2</sub>, median [IQR] | 0.65 [0.40-0.93] | 0.50 [0.40-0.95] |  |
| Time from birth (s) to |  |  |  |
| - First cry, median [IQR] | 65 [45-102] | 48[30-68] | 17 [5-31] |
| - Regular cries, median [IQR] | 88 [56-130] | 75 [52-100] | 29 [15-43] |
| Apgar score at |  |  |  |
| - 1 min, median [IQR] | 6 [5, 8] | 6 [5, 8] | 9 [9-9] |
| - 5 min, median [IQR] | 9 [9, 9] | 9 [9, 9] | 9 [9-9] |
| Cord arterial pH, median [IQR] | 7.21 [7.13-7.26];<br>n=12 | 7.14 [7.09-7.21];<br>n=13 | 7.23 [7.17-7.31]; n=42 |
| Cord venous pH, median [IQR] | 7.28 [7.24-7.34];<br>n=19 | 7.32[7.28-7.36];<br>n=15 | 7.29 [7.26-7.31]; n=66 |
| Infants with heart rate <100 bpm | 11 (37%) | 10 (40%) | 6 (6%) |
| Admitted for respiratory support | 1 (3%) | 2 (8%) | 1 (1%) |

<sup>a</sup> Includes hypertensive disorders of pregnancy, diabetes mellitus, sepsis, oligohydramnios, antepartum haemorrhage, placenta praevia.
<sup>b</sup> Prolonged 2<sup>nd</sup> stage: ≥3 hours (with epidural) and ≥2 hours (without epidural) in primiparae, and ≥2 hours (with epidural) and ≥1 hour (without epidural) in multiparae.
Abbreviations: bpm, beats per minute; CPAP, continuous positive airway pressure; CTG, cardiotocography; ECC, early cord clamping; IM, intramuscular; IQR, interquartile range; IV, intravenous; PBCC, physiologically-based cord clamping; SD, standard deviation.

Cord clamping occurred at a mean of 136 s and 36 s in the PBCC and ECC arms, respectively. All infants in the PBCC arm received the intended intervention. One infant in the ECC arm received DCC at clinician discretion. A larger proportion of infants in the PBCC group received any resuscitation (100% vs 76%, risk difference 24%, 95% confidence interval 7–41%).

### Primary outcome

The mean (SD) BP at 3-4 min after birth in the PBCC group was 64 mmHg (10 mmHg) compared to 62 mmHg (10 mmHg) in the ECC group (Table 2), mean difference 2 mmHg (95% confidence interval −3 – 8 mmHg, p= 0.42). These values were comparable to the 50^th^-75^th^ percentile for mean BP in non-randomised infants (60-69 mmHg). Mean BP was similar in the subgroups of emergency and non-emergency birth (≥ 36^+0^ weeks’ gestation); there was insufficient data for comparison in the preterm subgroup.

**Table 2.** Primary outcome and subgroup analyses.

|  | **PBCC arm**<br>**n = 30<sup>a</sup>** | **ECC arm**<br>**n = 25** | **Mean Difference**<br>**(95% CI)** | **p-**<br>**value** |
| --- | --- | --- | --- | --- |
| **Primary Outcome** |  |  |  |  |
| Mean Blood Pressure 3-4 min after birth<sup>b</sup> | 64 (10);<br>n= 25 | 62 (10)<br>n= 25 | 2 (-3, 8) | 0.42<sup>*</sup> |
| **Subgroup** |  |  |  |  |
| - 32<sup>+0</sup> to 35<sup>+6</sup> weeks' gestation | N/A;<br>n= 0 | 64 (14);<br>n= 3 | N/A |  |
| - Emergency birth ≥ 36<sup>+0</sup> weeks' gestation | 64 (12);<br>n= 15 | 63 (11);<br>n= 11 | -1 (-10, 8) |  |
| - Non-emergency birth ≥ 36<sup>+0</sup> weeks' gestation | 65 (9);<br>n= 10 | 64 (7);<br>n= 11 | -4 (-12, 3) |  |

<sup>a</sup> 5 infants in the PBCC arm had missing data for the primary outcome- 2 were in the 32<sup>+0</sup> to 35<sup>+6</sup> weeks' gestation subgroup, 2 were in the emergency birth ≥ 36<sup>+0</sup> weeks' gestation subgroup, and 1 in the non-emergency birth ≥ 36<sup>+0</sup> weeks' gestation subgroup.
<sup>b</sup> Values are mean (standard deviation) for each group.
<sup>\*</sup> p-value calculated from independent samples t-test.
Abbreviations: ECC, early cord clamping; PBCC, physiologically-based cord clamping.

### Secondary outcomes

There were no differences in systolic, mean or diastolic BP between randomised study arms at either the 3-4 min or 6-7 min timepoints (Figure 2). BP among infants in each randomised arm was similar between the 2 timepoints and approximated to the 25^th^–75^th^ percentiles of BP derived from infants in the observational arm.

**Fig 2.**
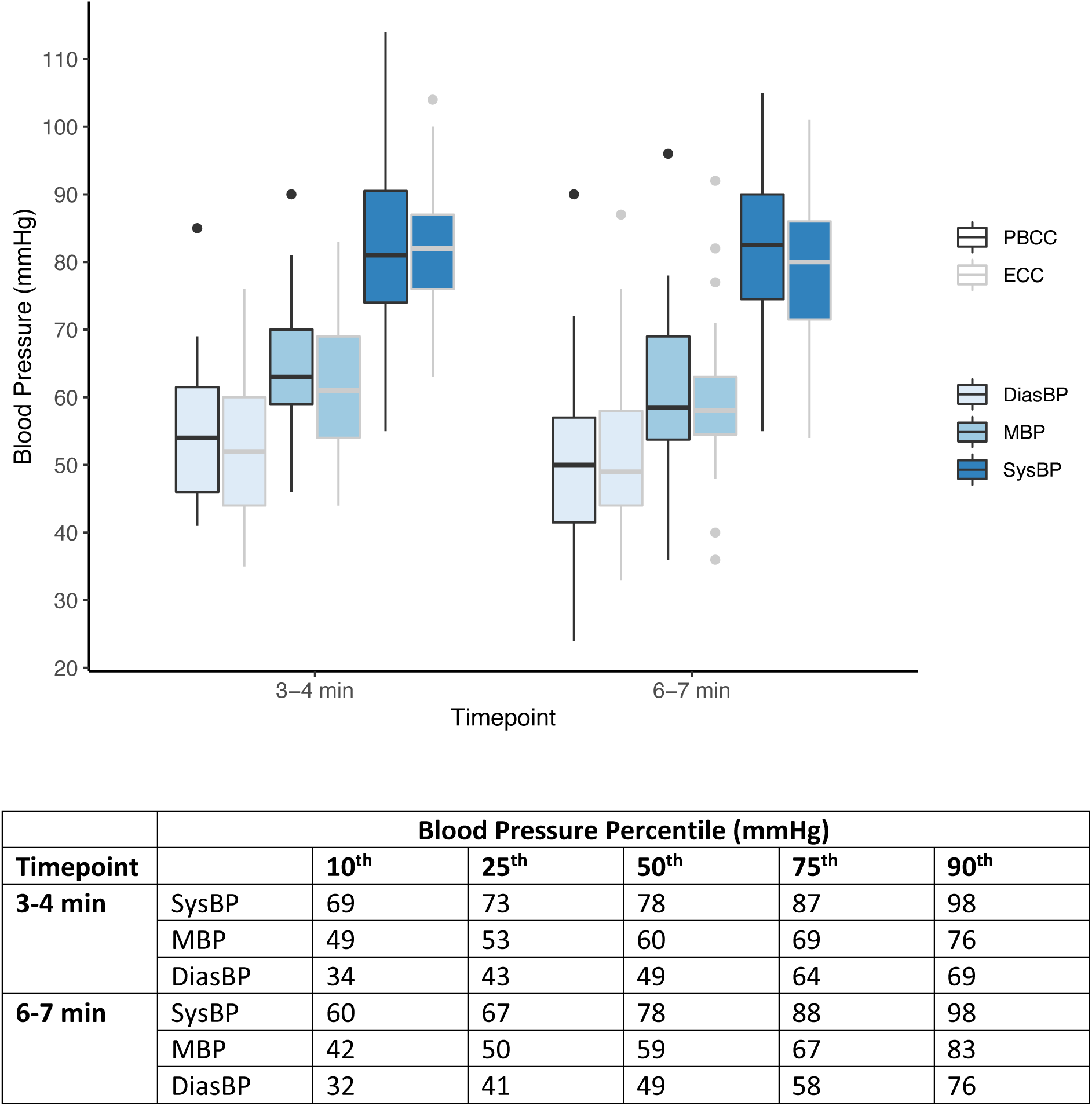
**(A)** Boxplots showing the median, interquartile range, and range of pre-ductal blood pressure (BP) in the physiologically-based cord clamping (PBCC) and early cord clamping (ECC) groups at the specified timepoints. **(B)** Blood pressure percentiles from non-randomised infants born at ≥35^+0^ weeks’ gestation who received deferred cord clamping. DiasBP, diastolic blood pressure; MBP, mean blood pressure; SysBP, systolic blood pressure.

Cerebral oxygenation and cFTOE was also similar between the randomised arms during the first 10 min after birth (Figure 3). At 1h after birth, the mean (SD) rStO_2_ was 77% (3%) in the PBCC arm (n=17) and 77% (4%) in the ECC arm (n=14). The mean (SD) rStO_2_ at 1h in the observational arm was 78% (4%) from n=36 infants. We were not able to acquire data in the majority of infants at the 1h timepoint because of infant breastfeeding or researcher non-availability.

**Fig 3.**
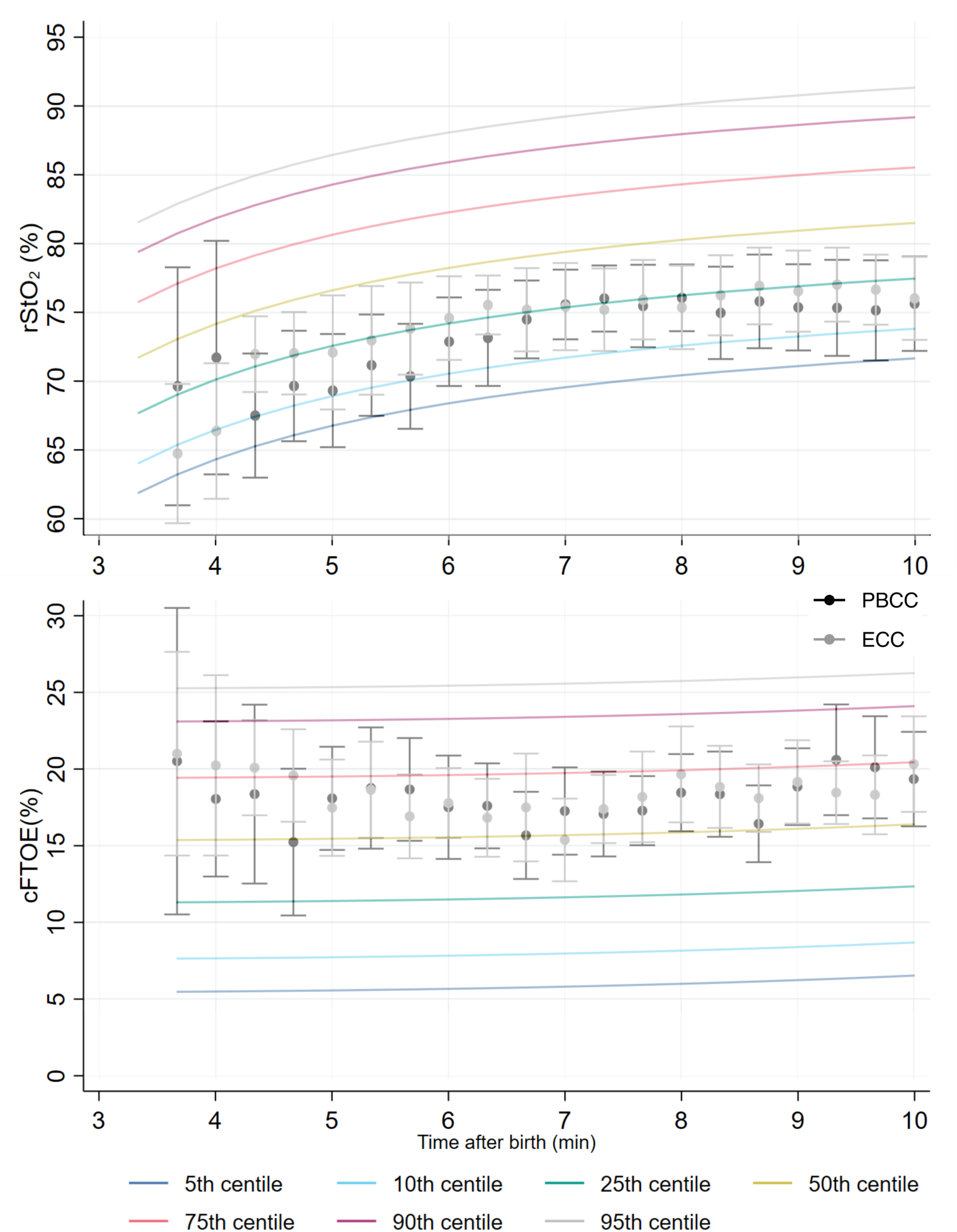
The 5th, 10th, 25th, 50th, 75th, 90th, and 95th percentiles for cerebral tissue oxygen saturation (rStO_2_) and fractional tissue oxygen extraction (cFTOE) for non-randomised infants born at ≥35^+0^ weeks’ gestation who received deferred cord clamping. The mean +/- 95% confidence intervals for rStO_2_ and cFTOE for the randomised infants who received either physiologically based cord clamping (PBCC) or early cord clamping (ECC) are also shown.

Percentiles for rStO_2_ and cFTOE are shown in Figure 3 and in the Supplement. Percentile charts subdivided by mode of birth are provided in the Supplemental Figures 3 & 6. rStO_2_ was lower but cFTOE similar in infants born by caesarean section (n= 49) compared to infants born vaginally (n= 57).

## Discussion

In this sub-study nested within an RCT, we did not observe differences in BP (at 3-4 minutes and 6-7 minutes) or cerebral oxygenation for infants ≥32^+0^ weeks’ gestation who received PBCC compared to ECC. These findings must be interpreted in the context of closely monitored births where infants required limited resuscitation, delivered by trained providers.

A previous study in moderately asphyxiated lambs found that PBCC protected against an overshoot in systemic BP after ventilation onset and associated cerebrovascular protein leakage.^2^ However, cord clamping in the PBCC group was 15 minutes after ventilation onset-much longer than was felt to be acceptable to clinicians when designing this trial. It is possible that a longer duration of PBCC in a more asphyxiated study population requiring advanced resuscitation would have a significant impact on BP and/or cerebral oxygenation when compared to ECC. The 3-4 min timepoint for our primary outcome was ∼3 min and ∼1 min after cord clamping in the ECC and PBCC groups respectively. In preterm lamb studies, there were similarly no BP differences at corresponding timepoints.^4,5^

With regards to cerebral oxygenation in the delivery room, a recent RCT found no differences between PBCC and ECC in vaginally-born, full-term neonates not needing resuscitation.^27^ Our findings were similar in a population that received resuscitation and included caesarean births. In contrast, Katheria et al. found that at 12 hours after birth, cerebral oxygen saturation and BP were higher among term infants at risk of needing resuscitation who were randomised to UCC at 5 min versus within 1 min after birth.^28^

This study provided an opportunity to concurrently describe the normal course of BP and cerebral oxygenation during unassisted transition with DCC at both caesarean and vaginal births. Our observational cohort is representative of births where physiological monitoring is likely to be used, i.e., when there is fetal or anticipated neonatal compromise. It may therefore be more appropriate for reference physiological percentiles to be derived from the population of at-risk births than from low-risk births where a paediatric clinician does not typically attend.

Pichler et al. and Salihoğlu et al. have previously reported BP percentiles for term infants shortly after birth.^22,23^ When comparing corresponding time points, the measurements obtained in our study were higher by approximately 10-15 mmHg. There are important differences between the studies that may explain this difference. Both previous studies included low-risk births and infants who received ECC. Pichler et al. measured cuff BP from the left upper arm. In contrast, we describe pre-ductal BP percentiles in a cohort of at-risk term births under conditions that would induce fetal stress, following DCC. Corresponding with this, the HR percentiles we recently reported from the Baby-DUCC cohort were substantially higher than those previously described by Dawson et al., which were derived from low-risk births with ECC.^17,19,29^

Study characteristics may explain, to some extent, the marked differences in cerebral oxygenation percentiles in comparison to previously published work. In a cohort of low-risk term births with ECC, Pichler et al. described the 10^th^ percentile for cerebral oxygenation to rise from around 45% at 5 min to 65% at 10 min.^14^ The corresponding values in our cohort were 68% rising to 74%. At the upper percentiles, cerebral oxygenation values are more comparable. Pichler et al. reported the 90^th^ percentile for cerebral oxygenation to rise from around 85% at 5 min to 90% at 10 min after birth. The corresponding values in our cohort were 84% rising to 89%. Pichler et al. used the Invos 5100 (Somanetics Corp, Troy, Michigan) system for measurements.

Similar to Pichler et al. we noted lower overall cerebral oxygenation values at caesarean births in comparison to vaginal births over the first 10 min (Supplemental Figure 3). Baik et al. reported cerebral oxygenation values for low-risk term caesarean births with ECC, measured using the NIRO 200NX (NIRO, Hamamatsu, Japan).^15^ Their 10^th^ percentile values were approximately 15% lower than ours between 5-10 min after birth, while 90^th^ percentile values were approximately 5% lower across the same timeframe. Corresponding with this pattern, our cFTOE values were substantially different at the higher percentiles (approximately 15-20% lower in comparison to Pichler et al. and Baik et al.) but more comparable at the lower percentiles.

Differences in measured cerebral oxygenation between NIRS devices are well described.^30^ While reference percentiles should ideally be device-specific, the cerebral oxygenation trend for an individual participant is likely to be most informative. Nevertheless, across studies, the 90^th^ percentile for cerebral oxygenation was found to be <90%. This cut-off may therefore be useful to define cerebral hyperoxia in near-term and term infants, for instance when studying oxygen supplementation after birth, particularly with cFTOE values <10% (10^th^-25^th^ percentile across studies).^6^ Given the marked variability of cerebral oxygenation values between studies at the lower percentiles, determining cut-off values for cerebral hypoxia is unlikely to be meaningful.

The strengths of this study include the methodological design that allowed randomisation to be rapidly performed once the infant was assessed as needing resuscitation. Over 20% of the randomised infants were enrolled via deferred consent, ensuring that we included infants with fetal compromise significant enough to require emergency birth. Adherence to study interventions was high and physiological outcomes were assessed blind to randomised group allocation. For the creation of the percentile charts, we used statistical methodology that accounts for the variation in repeated measurements for each infant over time.^24^ The major limitation was that we were unable to recruit a cohort of infants requiring prolonged or advanced resuscitation. Trial participation may have improved the ability of clinicians to intervene early before infants in either arm became significantly compromised. We did not measure BP and cerebral saturation at time points after the delivery room. Though adequately powered, our sample size was relatively small. We were therefore unable to correlate our physiological measurements with important clinical outcomes like neonatal unit admission.

## Conclusions

In this study, we found no evidence of a difference in BP (at 3-4 minutes and 6-7 minutes) or cerebral oxygenation with 2 different cord clamping strategies for late preterm and term infants receiving resuscitation at birth. The percentiles for both BP and cerebral oxygenation represent the first, to our knowledge, for late preterm and term infants receiving DCC, in line with contemporary recommendations.

## Data availability

The datasets generated and analysed during the current study are available from the corresponding author on reasonable request.

## Acknowledgements

None

## Author contributions

Substantial contributions to conception and design: SB, DAB, FYW, CTR, SBH, GRP, PGD. Substantial contributions to acquisition of data: SB, DAB. Substantial contributions to analysis and interpretation of data: all authors. Drafting the article or revising it critically for important intellectual content: all authors. Final approval of the version to be published: all authors.

## Consent statement

Written consent for participation in this sub-study was obtained by the parents prior to the infant’s birth in addition to consent for participation in the parent (Baby-DUCC) trial.

