## Supplementary material for "Blood Pressure and Cerebral Oxygenation with Physiologically-based Cord Clamping: A Sub-Study of the BabyDUCC Trial"

**Supplemental Figure 1.** The 5th, 10th, 25th, 50th, 75th, 90th, and 95th percentiles for cerebral tissue oxygen saturation ($rStO_2$) for non-randomised infants born vaginally or by caesarean section at ≥$35^{+0}$ weeks' gestation who received deferred cord clamping.

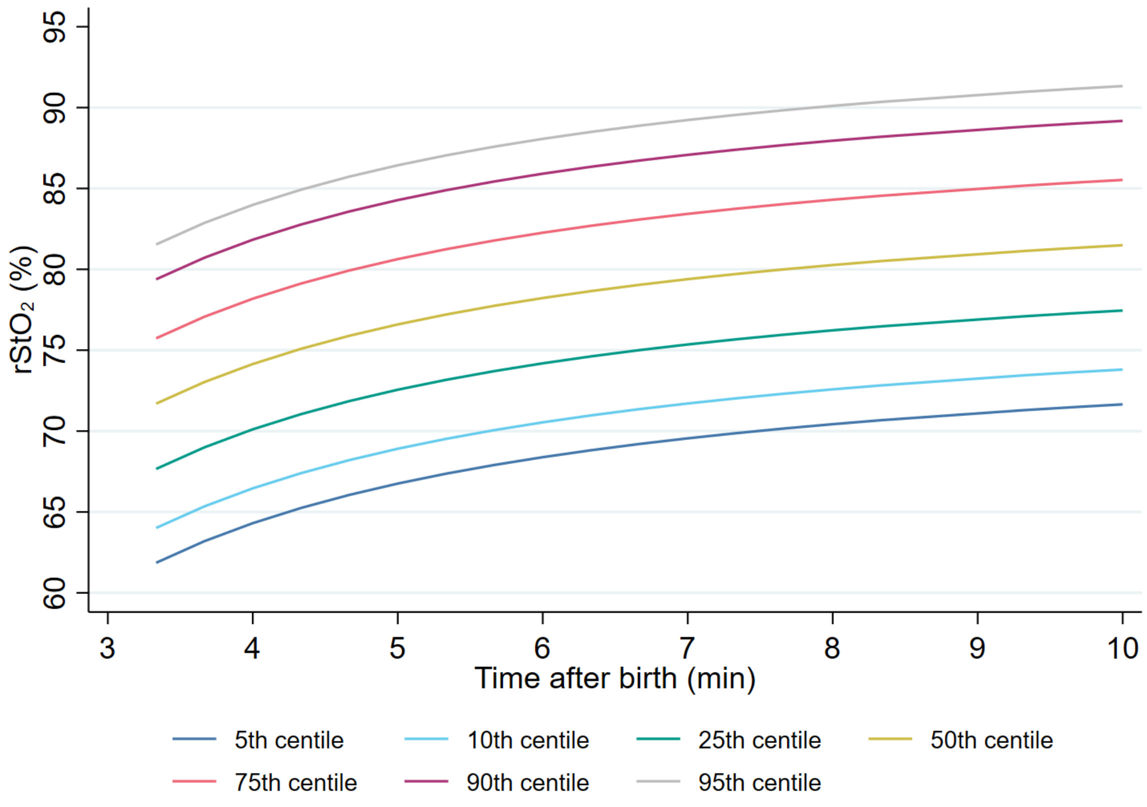

**Supplemental Figure 2.** Individual trajectories of cerebral tissue oxygen saturation ($rStO_2$) among non-randomised infants born vaginally or by caesarean section at ≥35$^{+0}$ weeks' gestation who received deferred cord clamping.

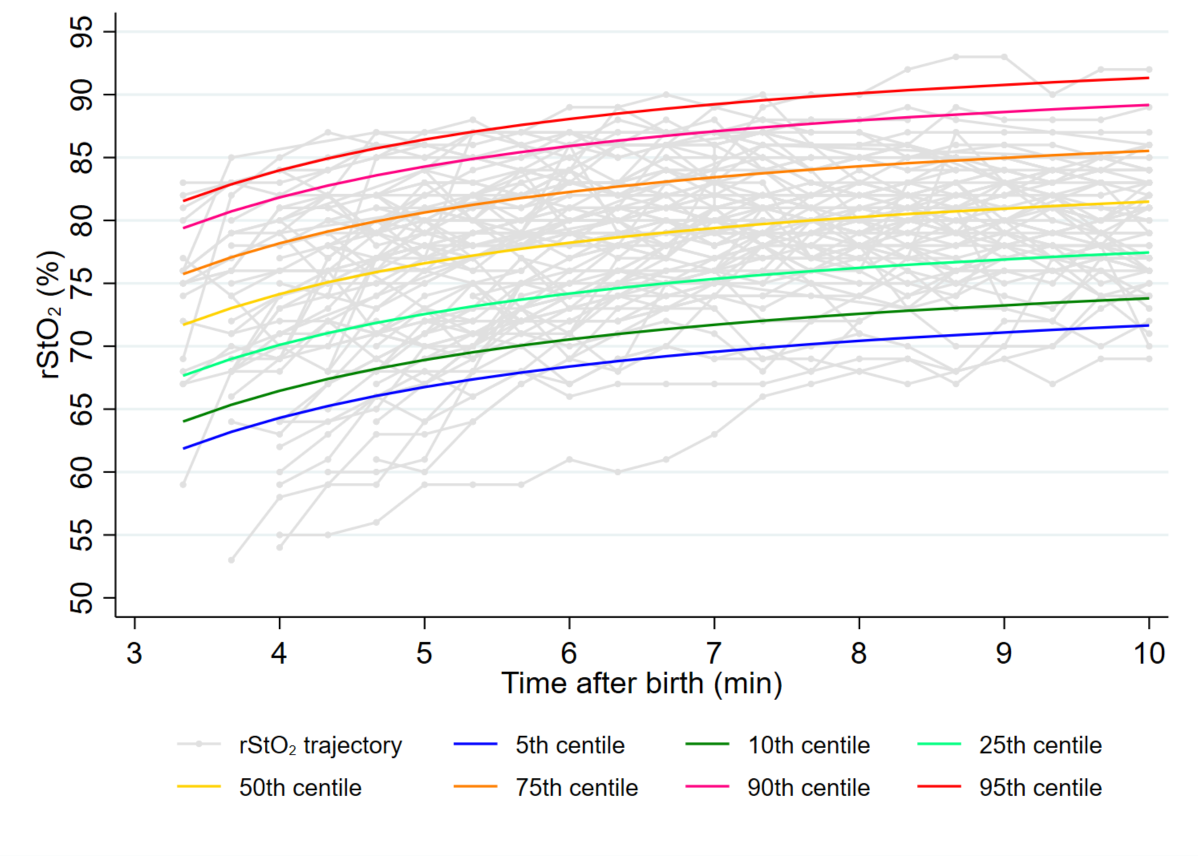

**Supplemental Figure 3.** The 5th, 10th, 25th, 50th, 75th, 90th, and 95th percentiles for cerebral tissue oxygen saturation ($rStO_2$) for non-randomised infants born by caesarean section (left) and vaginal birth (right) at ≥35$^{+0}$ weeks' gestation who received deferred cord clamping.

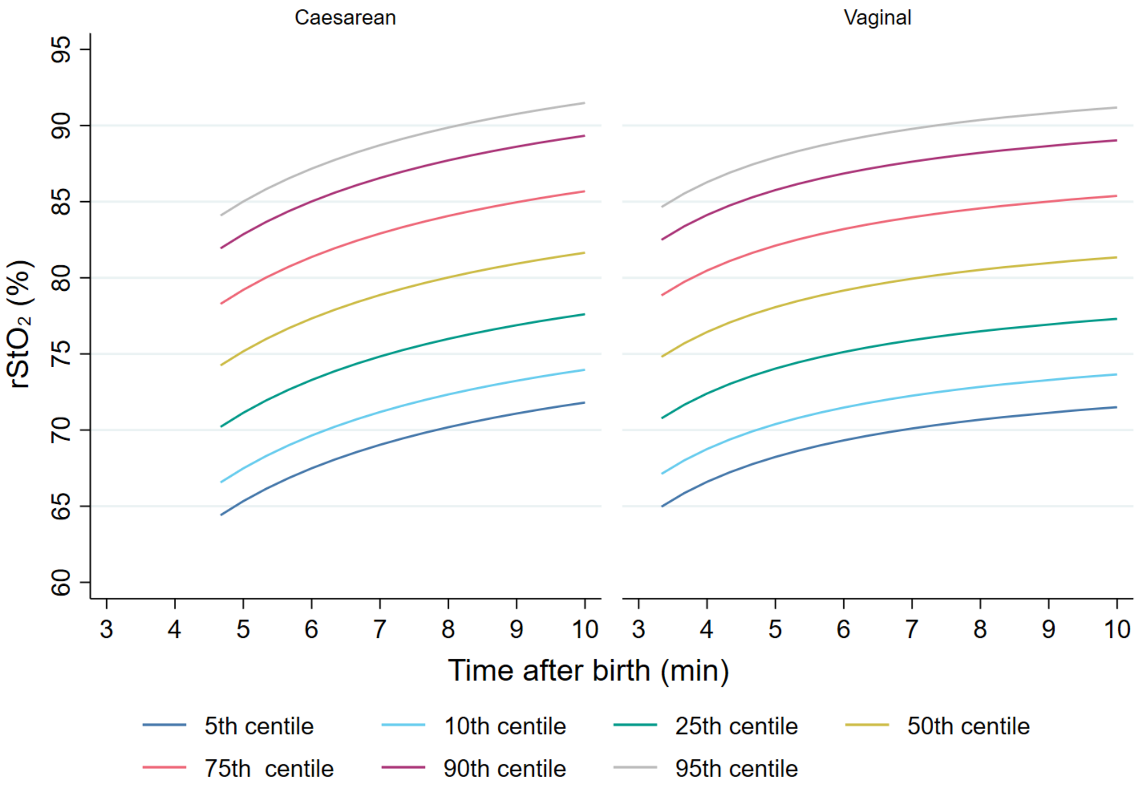

**Supplemental Figure 4.** The 5th, 10th, 25th, 50th, 75th, 90th, and 95th percentiles for cerebral fractional tissue oxygen extraction (cFTOE) for non-randomised infants born vaginally or by caesarean section at ≥35$^{+0}$ weeks' gestation who received deferred cord clamping.

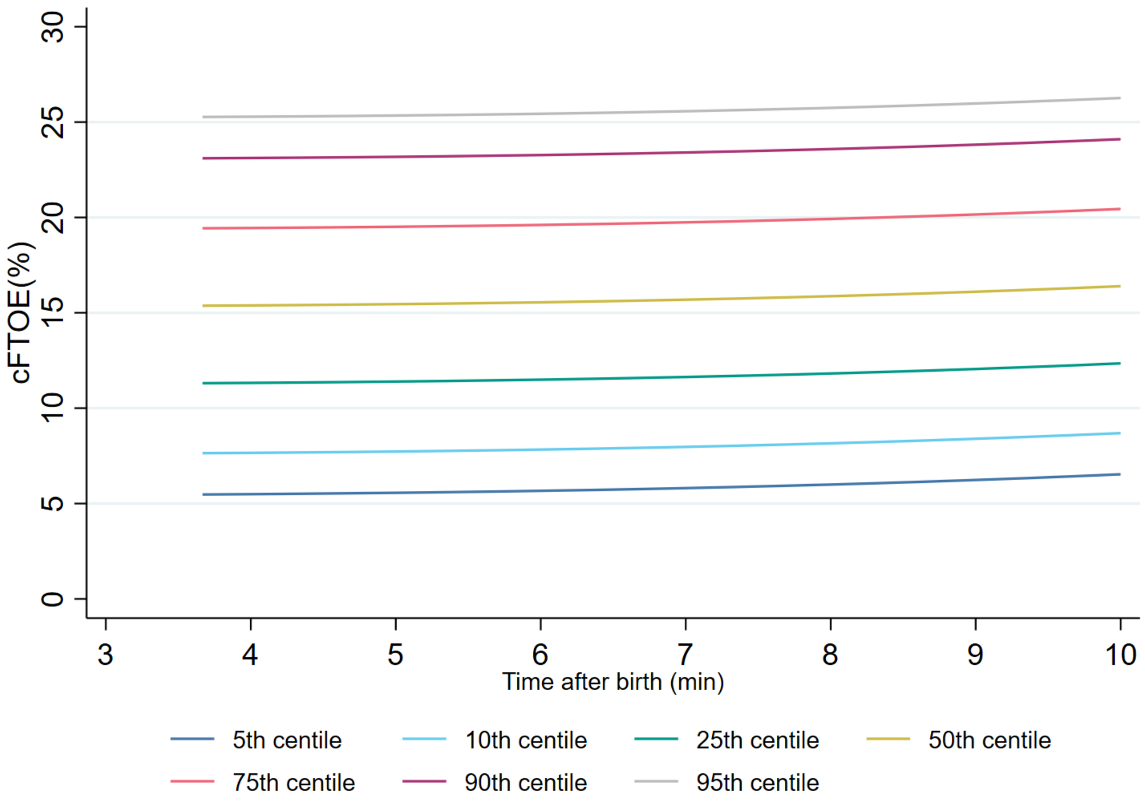

**Supplemental Figure 5.** Individual trajectories of cerebral fractional tissue oxygen extraction (cFTOE) among non-randomised infants born vaginally or by caesarean section at ≥35$^{+0}$ weeks' gestation who received deferred cord clamping.

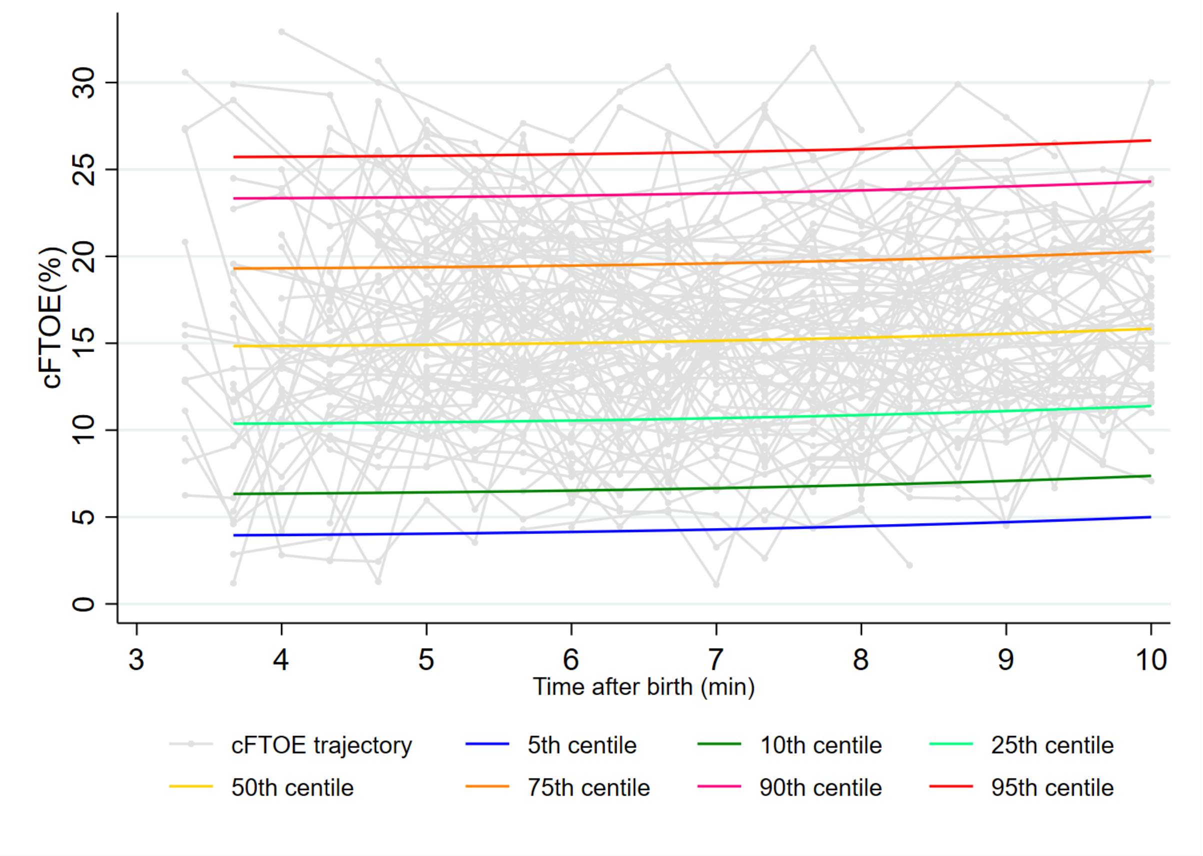

**Supplemental Figure 6.** The 5th, 10th, 25th, 50th, 75th, 90th, and 95th percentiles for cerebral fractional tissue oxygen extraction (cFTOE) for non-randomised infants born by caesarean section (left) and vaginal birth (right) at ≥35$^{+0}$ weeks' gestation who received deferred cord clamping.

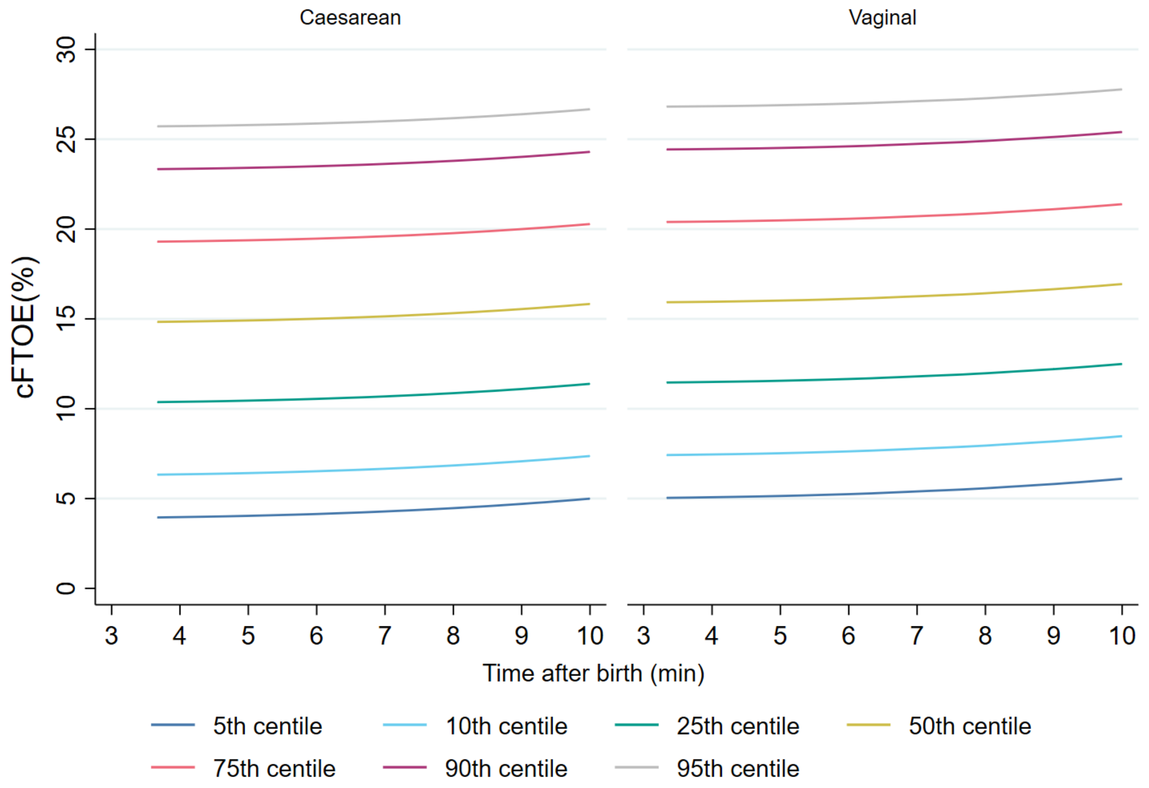
